# Towards Longitudinal Glioma Segmentation: Evaluating combined pre- and post-treatment MRI training data for automated tumor segmentation using nnU-Net

**DOI:** 10.1101/2023.05.31.23290537

**Authors:** Sara Ranjbar, Kyle W. Singleton, Lee Curtin, Lisa Paulson, Kamala Clark-Swanson, Andrea Hawkins-Daarud, J. Ross Mitchell, Pamela R. Jackson, Kristin R. Swanson

**Author notes:** Authors contributed equally (Pamela R. Jackson, Kristin R. Swanson).

## Abstract

Identification of key phenotypic regions such as necrosis, contrast enhancement, and edema on magnetic resonance imaging (MRI) is important for understanding disease evolution and treatment response in patients with glioma. Manual delineation is time intensive and not feasible for a clinical workflow. Automating phenotypic region segmentation overcomes many issues with manual segmentation, however, current glioma segmentation datasets focus on pre-treatment, diagnostic scans, where treatment effects and surgical cavities are not present. Thus, existing automatic segmentation models are not applicable to post-treatment imaging that is used for longitudinal evaluation of care. Here, we present a comparison of three-dimensional convolutional neural networks (nnU-Net architecture) trained on large temporally defined pre-treatment, post-treatment, and mixed cohorts. We used a total of 1563 imaging timepoints from 854 patients curated from 13 different institutions as well as diverse public data sets to understand the capabilities and limitations of automatic segmentation on glioma images with different phenotypic and treatment appearance. We assessed the performance of models using Dice coefficients on test cases from each group comparing predictions with manual segmentations generated by trained technicians. We demonstrate that training a combined model can be as effective as models trained on just one temporal group. The results highlight the importance of a diverse training set, that includes images from the course of disease and with effects from treatment, in the creation of a model that can accurately segment glioma MRIs at multiple treatment time points.

## 1 Introduction

Gliomas are highly invasive primary brain malignancies. The most aggressive form, glioblastoma, has a dismal median survival of ∼15 months, despite extensive treatment [1]. As their eloquent location limits longitudinal access to the tumor tissue, pragmatic insights into tumor changes are inferred through the lens of clinical imaging, particularly magnetic resonance imaging (MRI). Clinical imaging and accurate segmentations are central to the modern management of glioma and are crucial to advancing personalized medicine. Ideally, treatment planning and response evaluation for gliomas would be based on accurate segmentations from MRI, resulting in quantitative measurements of tumor size, measured across the full timeline of patient care. However, in clinical practice, subjective summary assessments are typically made to ascertain disease progression/regression [2]. Clinicians often adopt more granular measures because manual glioma segmentation is an extraordinarily time-consuming and visually difficult task where variability can be difficult to minimize between even highly trained observers.

When evaluating post-treatment brain scans, tumor segmentation is demonstrably more challenging than the pre-treatment setting. This is due to the added effects that surgery, radiation, chemotherapy, and immunotherapy can have on brain appearance including surgically imposed cavities, treatment-induced inflammation, and radiation necrosis. For example, a resection cavity containing a hematoma or cerebrospinal fluid (CSF) can be confounded with enhancing tumor on T1-weighted post contrast injection MRI (T1Gd) due to post-surgical blood degradation products [3] (see **Fig. 1**; Day 18). Another post-surgical challenge is that residual tumor tissue often appears as small enhancing volumes that are more difficult to measure with consistency while avoiding other image and therapy artifacts [4].

**Fig. 1.**
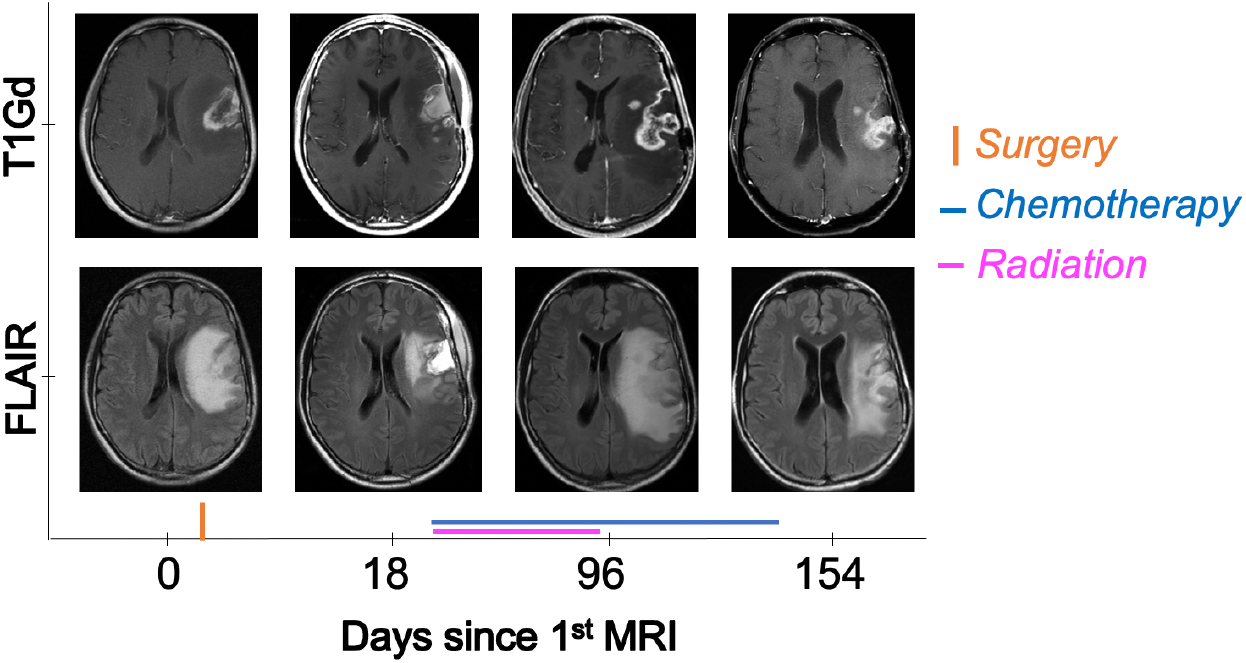
Dynamic change in tumor appearance throughout treatment. Images belong to a male patient in his 40s with a left lateral inferior frontal lobe glioblastoma. Hyperintense signal in T1Gd imaging (top) is gadolinium leakage through disrupted blood brain barrier, associated with the bulk of the tumor, with dark internal signal showing a necrotic core. Hyperintensity signal on T2/FLAIR imaging (bottom) reflects tumor-associated edema. The strikingly hyperintense FLAIR signal at 18 days is related to post-surgical effects in the resection cavity. Texture and intensity of the bright signal are variable across time in both MRI sequences.

Although the literature on brain tumor segmentation is abundant, the published work and segmentation challenges tend to focus on pre-treatment data because of the difficulty and uncertainty in the post-treatment setting. To the best of our knowledge, no previous work has assessed the effect of combined pre- and post-treatment training data on the performance of glioma segmentation models. In this work, we address this gap in research to showcase the difference between performance of models trained and tested on pre-treatment, post-treatment, and mixed data and applied to data from each scenario.

## 2 Materials and Methods

### 2.1 Study Design, Patients, and Imaging Data

This study was approved by the institutional review board of the main data collection site and followed the Health Insurance Portability and Accountability Act. The internal repository was searched for adult patients with a diagnosis of glioma of any grade, with available 3D post gadolinium contrast injection T1-weighted images (T1Gd) and T2 fluid-attenuated inversion recovery (FLAIR) imaging as well as manual tumor annotation. Cases with a low-quality of brain mask (see brain mask generation process described in section 2.2) were excluded from the analysis. The final cohort included imaging sets from 1563 imaging timepoints from 854 patients (mean age, 53 years ±14 (SD), 51.4% men) evaluated between 1990 and 2021 across 13 institutions and from the TCGA and TCIA public datasets. Patients had a range of diagnoses (610 glioblastoma Grade IV, 57 Grade III, 63 low-grade glioma, 25 metastases, and 99 Unknown) and had undergone various treatment plans typical for glioma patients including gross total or subtotal tumor resection, rounds of radiation and chemotherapy. No treatment-related exclusion criterion was applied to the schedule or type of therapy to allow for the full diversity of treatment effects present on MRI. Similarly, we did not select cases based on whole tumor or region size, as presentation varies widely across grades and post-treatment MRI abnormalities are often smaller in size due to therapy (particularly for enhancing tumor).

Given the range of dates and the number of institutions included in the study, image acquisition protocols were diverse and span across a wide variety of scanner manufacturers, field strengths (both 1.5T and 3T), repetition times, echo times, field of view, matrix size, and slice thickness. We did not exclude any imaging based on image acquisition protocol.

Image sets with available manual labels were split into train and test cohorts using a 70-30 ratio (train:1090, test:473) on the axis of patient, ensuring that all scans from a given patient were assigned to either the train or test cohort to avoid data leakage. No additional biological or demographic variables were considered in the inclusion or exclusion criteria.

### 2.2 Image Preprocessing

We implemented an automated image processing pipeline which included co-registration of T1Gd and FLAIR images using the elastix toolbox [5], 1 × 1 × 2mm interpolation, and zero padding or cropping to a 280 × 280 × 112 image dimension to resize images. Next, denoising with ITK’s CurvatureFlow algorithm, N4 intensity bias field correction [6], and z-score intensity normalization (zero mean and unit standard deviation) were applied. Finally, each image was skull-stripped using the union of SPM12 [7] tissue probability maps, following the process described in Ranjbar et al. [8]. The final brain masks were reviewed by a trained individual prior to their use for skull stripping and timepoints with a low quality of brain mask were excluded from the analysis.

### 2.3 Reference Tumor Annotations

The reference tumor segmentations were generated by a team of junior technicians trained and refereed by an experienced technician. Annotations included contrast enhancing tumor (CE), necrotic core (NEC), and edema (ED). In brief, contrast enhancement (CE) is defined as hyperintense regions on T1Gd images, necrotic (NEC) core is defined as the hypointense areas within the CE region and hyperintensity on FLAIR is defined as edema (ED) [9]. The union of all 3 labels was defined as the whole tumor (WT). We also used the union of CE and NEC (CE+NEC), as these are the abnormalities present on T1Gd MRI.

### 2.4 Convolutional Neural Network Architecture and Training

The 3D full resolution nnU-Net [10] was utilized for model training with default settings. The nnU-Net is a self-configuring framework that has shown remarkable success in previous medical segmentation tasks including in previous BraTS challenges [11]. We used the default setting of nnU-Net. This included training in a 5-fold cross validation scheme with a fixed 1000 epochs, 250 iterations, and batch size of 2. Details of the optimization method were left at default, which included using an SGD optimizer with initial learning rate of 0.01 and momentum of 0.99.

We conducted three rounds of training with this architecture and setting, changing the training cohort each time. First, we used only pre-treatment training cases (N=502) which is referred to as the ‘Pre’ model in the subsequent sections. Second, we conducted model training with only post-treatment training cases (N=588). This model is referred to as the ‘Post’ model here on. Finally, we merged the pre- and post-treatment cohorts and trained with the combined cases (N=1090). This model is referred to as the ‘Mixed’ model in the following section. All models were validated against previously unseen test cases from pre-treatment (N=219) and post-treatment (N=254).

### 2.5 Performance Metrics and Statistical Analysis

We report Dice coefficients of NEC, CE, CE+NEC, ED, and WT for the Pre, Post, and Mixed models on pre-treatment and post-treatment test cases. The statistical significance between models was assessed on each label region using within-subjects ANOVA analyses, with post-hoc pairwise Wilcoxon tests. The type I error rate was set at 0.05 after Bonferroni correction.

## 3 Results

**Table 1** presents the mean and standard deviation of Dice coefficients of the Pre, Post, and Mixed models when applied to pre- and post-treatment test cohorts. The boxplot version of these scores is presented in **Fig. 2**. As expected, the Pre model outperformed the Post model on pre-treatment test data, and the Post model outperformed the Pre model on post-treatment test data (p-values <0.0001, with moderate to large effect sizes between 0.31 and 0.85). On pre-treatment data, the average Dice coefficients for Pre and Mixed models were quite similar (**Fig. 3**, top). The Pre model outperformed the Mixed model in segmenting ED and WT (both p-values <0.0001, moderate effect sizes of 0.30 and 0.33 respectively), but not in NEC, CE or CE+NEC regions.

**Table 1.** Comparison of Models in pre- and post-treatment data Model Test data Mean±Std of Dice Coefficient per label

| Model | Test data | Mean±Std of Dice Coefficient per label |  |  |  |  |
| --- | --- | --- | --- | --- | --- | --- |
|  |  | ED | CE | NEC | CE+NEC | WT |
| Pre | Pre<br>N=219 | 0.78±0.13 | 0.80±0.14 | 0.53±0.29 | 0.81±0.16 | 0.86±0.11 |
| Post |  | 0.71±0.18 | 0.75±0.17 | 0.46±0.28 | 0.76±0.19 | 0.79±0.15 |
| Mixed |  | 0.77±0.13 | 0.79±0.16 | 0.53±0.29 | 0.80±0.17 | 0.85±0.12 |
| Pre | Post<br>N=254 | 0.73±0.13 | 0.69±0.12 | 0.28±0.26 | 0.68±0.13 | 0.78±0.09 |
| Post |  | 0.75±0.14 | 0.75±0.12 | 0.36±0.25 | 0.74±0.13 | 0.80±0.09 |
| Mixed |  | 0.75±0.14 | 0.74±0.13 | 0.35±0.25 | 0.73±0.14 | 0.80±0.09 |

**Fig. 2.**
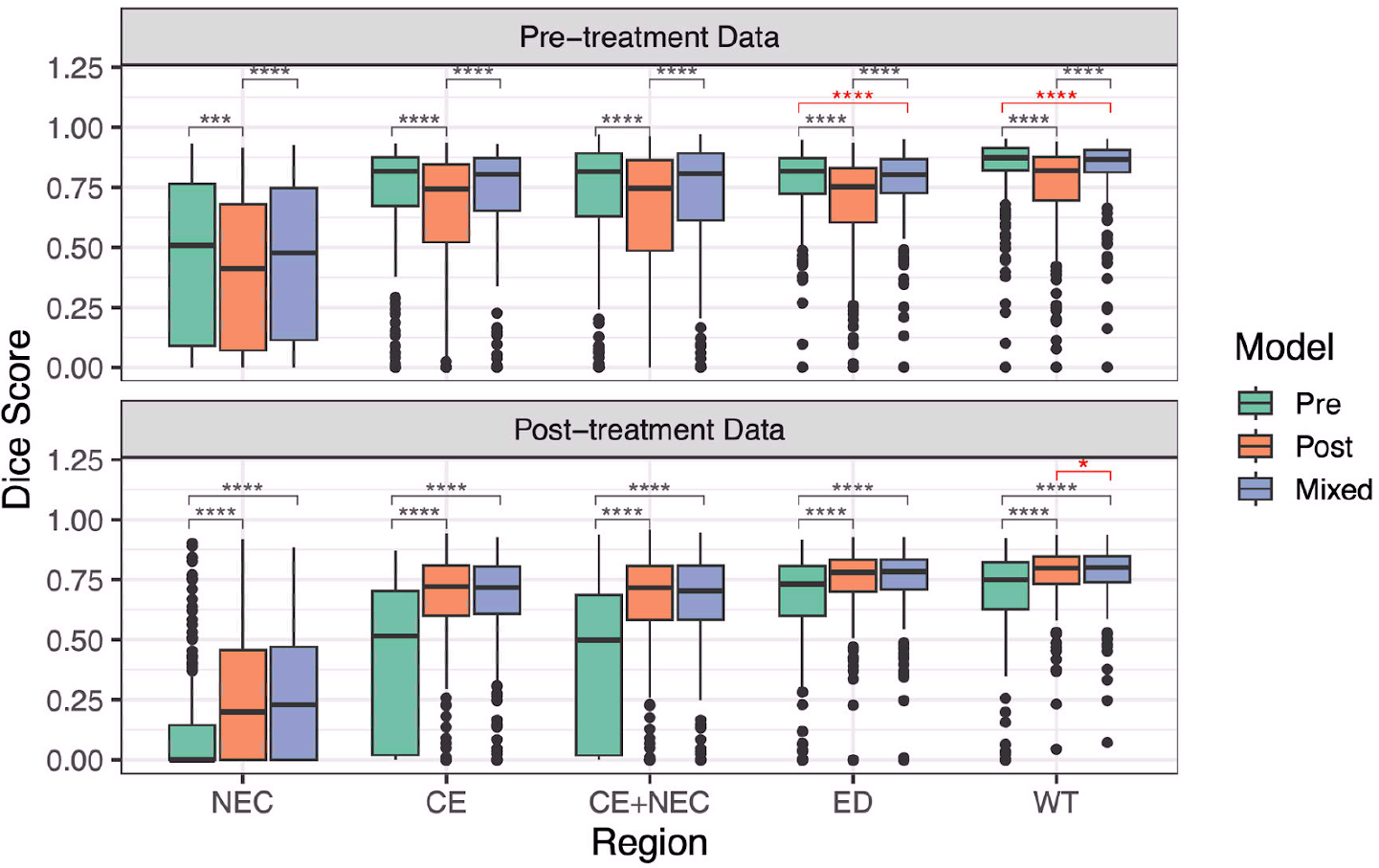
Comparison of model performance on different regions, on different test sets (*p<0.05, **p<0.01,***p<0.001,****p<0.0001; **red** indicates significant comparisons between the mixed model and the model trained and tested on the same type of data). As expected, the post-treatment model (**orange**) tested on pre-treatment data (top) consistently performs the worst, as does the post-treatment model (**green**) tested on pre-treatment data (bottom: all comparisons p<0.001 in gray). The mixed model (**blue**) consistently competes across both test sets with the model trained solely on the test set data. The mixed model was slightly outperformed by the pre-treatment model in the pre-treatment test case within CE+NEC (p<0.0001) and WT (p<0.0001) regions, while the mixed model slightly outperformed the post-treatment model in the post-treatment test case in WT regions (p=0.038).

**Fig. 3.**
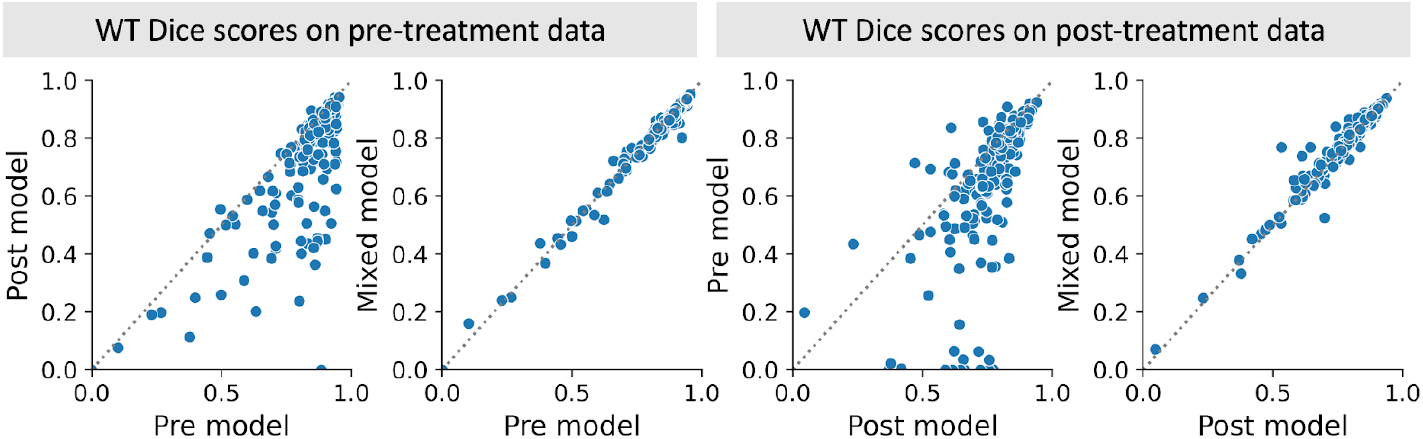
Comparison of Dice coefficients between pre-treatment, post-treatment, and mixed models for whole tumor (WT) segmentation. Dotted line indicates the line of identity. Overall, for pre-treatment test data, the pre-treatment model performs better whereas for the post-treatment test data, the post-treatment model performs better. Mixed model predictions are very similar to both pre- and post-treatment specific model predictions (96.8% and 95.6% of cases within 0.05 of each other respectively) while pre- and post-treatment models do not agree closely (only 56.0% and 46.9% of cases within 0.05 for pre- and post-treatment test data, respectively).

Similarly, in the post-treatment analysis, the Mixed model average Dice coefficients were comparable to the Post model (**Fig. 3**, bottom), while the Pre model underperformed in the same task. Interestingly, the Mixed model performed significantly better than the Post model in segmenting WT (p=0.038, a small effect size of 0.16). **Fig. 4** visualizes predictions of models in example pre- and post-treatment cases. As expected, not having previously seen cases from the other cohort, the pre-treatment model had some difficulty excluding the resection cavity (second example from the right) and the post-treatment model, in some cases, with the necrotic core.

**Fig. 4.**
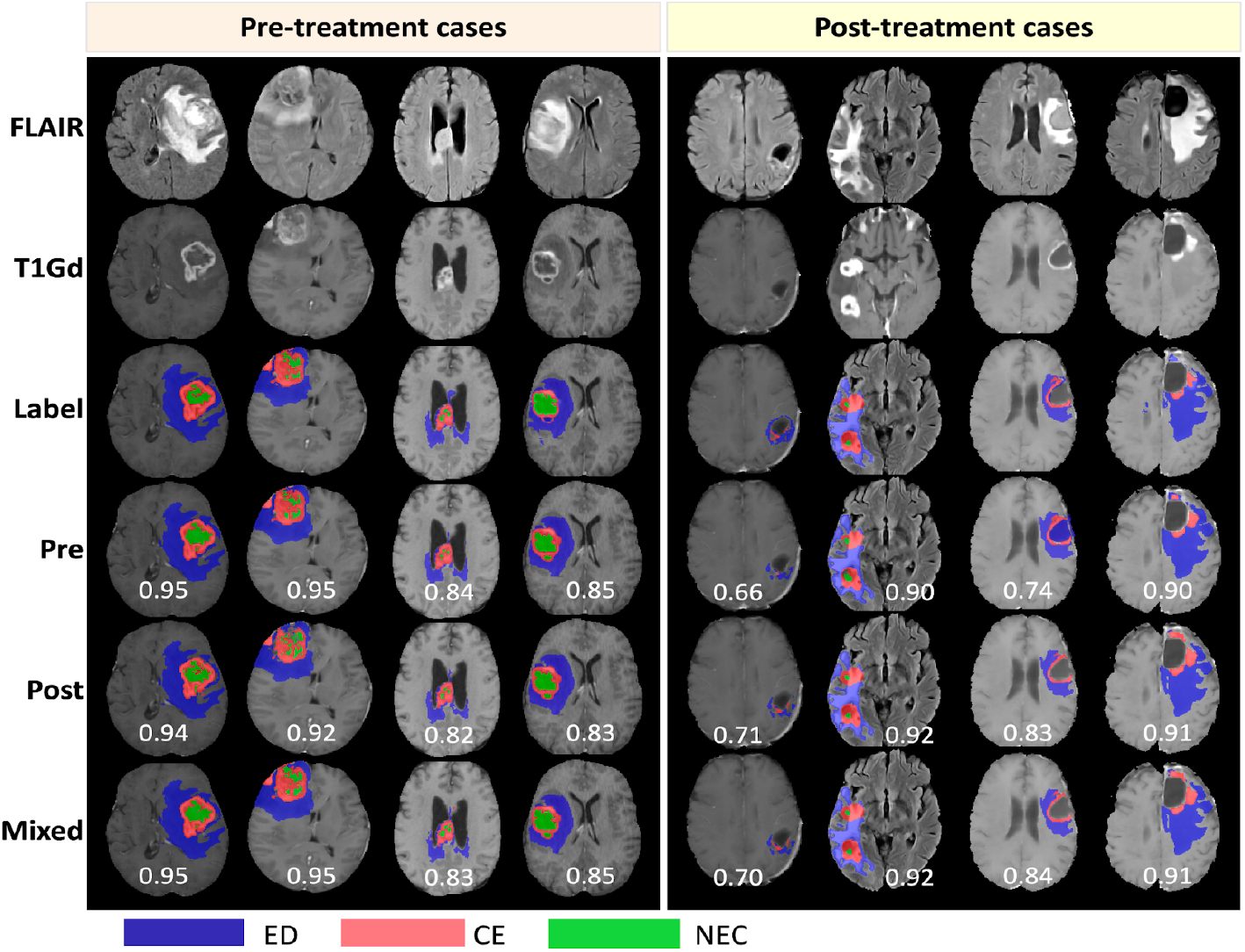
Examples of tumor segmentation predictions from each model on pre- and post-treatment data for the three regions of edema (ED - **blue**), contrast enhancement (CE - **red**), and necrotic core (NEC - **green**). Four example cases are shown for each time frame with input FLAIR and T1Gd images, the manually segmented labels, and predictions from each model. The numbers on prediction rows represent the whole tumor Dice score of the models.

## 4 Discussion

Few automatic segmentation papers, all of which aimed to measure longitudinal change in tumor burden to assess treatment response, have reported the performance of models in the post-treatment setting [12-14]. Rudie et al. [12] focused on proposing a solution for assessing change in tumor size (progressed vs not) by training a model on subtracted images in two consecutive timepoints to detect longitudinal change in a cohort of patients with diffuse gliomas. Although the context of their work was not directly com parable to ours, as part of their analysis the authors trained a baseline model on post-treatment images and reached mean Dice coefficients of 0.85 in Edema and 0.71 in active tumor regions.

Chang et al. [13] trained separate models for segmenting CE and ED in a small post-treatment glioma cohort and reached mean Dice coefficients of 0.70 for Edema, and 0.696 for CE. Kickingereder et al. [14] conducted a similar analysis but with a much larger sample size and reached a median Dice coefficient of 0.93 in non-enhancing tumor (equivalent to ED label) and 0.88 in CE. Our results are within the range of these works with mean Dice coefficients of 0.75 for edema, 0.73 for CE+NEC, and 0.80 for WT for the mixed model. It is largely understood that because of the various ways that treatment can affect patient images, a model trained solely on pre-treatment data will likely have decreased performance on post-treatment images. The result of the comparison between pre- and post-treatment models tested on data from the opposing cohort confirmed this assumption, as each model’s performance dropped drastically. Yet, our results also demonstrate that the combined information from both datasets can be trained together to provide a singular mixed model that produces comparable segmentation results to the specialized models.

One of the differences between our work and others is the number of input images included in our training data. The use of all four anatomical MRI sequences (T1, T1Gd, T2 and FLAIR) is common practice in the literature, especially as the commonly used BraTS challenge datasets offer all four. Previous works have demonstrated that using fewer sequences can result in comparable dice scores in glioma segmentation tasks [15]. We chose to include only two sequences because of the prevalence of these two sequence types in our clinical database, and the fact that in a clinical setting, all four sequences may not be incorporated in an imaging protocol or available for access. An alternative approach to excluding sequence types in favor of practicality could be training a sequence-agnostic model where the model is robust to missing sequences [16]. Including T2W images may improve the model’s ability to distinguish resection cavities. Besides increasing available information, T2W images generally have better signal to noise ratio and image quality which could improve the segmentation performance.

We recognize several limitations in this work. First, we use trained technician-generated labels for segmentations, rather than expert neuroradiologist-annotated labels. While it is possible these labels have reduced accuracy compared to experts, our technicians follow rules devised and described by our experts to try to maximize accuracy and minimize variability. The quality of training labels is a well-known factor in model performance and one that we would like to address further in future works. We took a minimal approach to our inclusion criteria apart from the patient age (>18) and availability of imaging and segmentation labels. As a result, while our data includes mostly examples from high- and low-grade glioma, some metastatic and unconfirmed cases are included in our work and treatments for patients were highly varied. These cases may impair model training and resulting segmentations in certain regions. Yet, we believe our segmentation accuracy demonstrates that segmentations can be robust while including this wide heterogeneity from many cases. Having a more selective data collection approach might explain higher performance of Kickingereder et al. [14] compared to others, showcasing the importance of quality training data for model training. Aggregating a rich dataset that more specifically tracks the broad range of possible phenotypic appearance of gliomas after treatment and ensuring a good representation of difficult cases could result in model improvement in the future.

To summarize, this work demonstrated that it is reasonable to combine pre- and post-treatment data for segmenting gliomas across all stages of treatment. Given that the majority of data in clinical practice follows the administration of therapy to the patient, this finding can help guide future work in incorporating this information for model training. In the future, we anticipate applying our mixed model for segmentation predictions that enable building a longitudinal glioma segmentation approach for response assessment.

## Data Availability

Due to the proprietary nature of patient data and patient information, we are not at liberty to freely share data with readers. However, data may be available for sharing upon the reasonable request of qualified parties as long as patient privacy and intellectual property interests of our institution are not compromised.

## Acknowledgements

The authors would like to thank the numerous members of our lab’s image analysis team that have given their time, attention, and talents over the years to generate regional segmentations. We are truly grateful to our funding sources: Mayo Clinic Marley Endowment Funds, NIH grants U01CA220378, U01CA250481, R01NS060752, R01CA164371, U54CA143970, U54CA193489, U54CA210180, the James S. McDonnell Foundation, and the Ben and Catherine Ivy Foundation.

